# SIM-CNN: Self-Supervised Individualized Multimodal Learning for Stress Prediction on Nurses Using Biosignals

**DOI:** 10.1101/2023.08.25.23294640

**Authors:** Sunmin Eom, Sunwoo Eom, Peter Washington

## Abstract

Precise stress recognition from biosignals is inherently challenging due to the heterogeneous nature of stress, individual physiological differences, and scarcity of labeled data. To address these issues, we developed SIM-CNN, a self-supervised learning (SSL) method for personalized stress-recognition models using multimodal biosignals. SIM-CNN involves training a multimodal 1D convolutional neural network (CNN) that leverages SSL to utilize massive unlabeled data, optimizing individual parameters and hyperparameters for precision health. SIM-CNN is evaluated on a real-world multimodal dataset collected from nurses that consists of 1,250 hours of biosignals, 83 hours of which are explicitly labeled with stress levels. SIM-CNN is pre-trained on the unlabeled biosignal data with next-step time series forecasting and fine-tuned on the labeled data for stress classification. Compared to SVMs and baseline CNNs with an identical architecture but without self-supervised pre-training, SIM-CNN shows clear improvements in the average AUC and accuracy, but a further examination of the data also suggests some intrinsic limitations of patient-specific stress recognition using biosignals recorded in the wild.

## 1. Introduction

Stress can negatively impact health, with prolonged exposure leading to pathological conditions including cognitive, immune, and cardiovascular abnormalities (Yaribeygi et al., 2017). The development of wearable devices that can recognize stress in real-time is a growing field of study (Samson & Koh, 2020) within precision health, an emerging discipline that focuses on individualized prevention strategies, diagnoses, and treatments (Silvera-Tawil et al., 2020).

Traditional stress-recognition methods, such as the cortisol test, require medical testing that is too burdensome for general use and is sometimes unavailable (Riley, 2012). General-purpose tests with physiological data do not account for variations across individuals. While machine learning (ML) is a natural solution for this task, most ML frameworks only utilize manually-annotated data that is difficult to obtain, especially in healthcare settings due to privacy and data transfer issues (Kim et al., 2019; Kruse et al., 2016; Miotto et al., 2017).

To accurately predict stress while addressing the intrinsic challenges associated with the multifaceted and subjective nature of stress data, we developed the self-supervised individualized multimodal convolutional neural network (SIM-CNN) framework. Our proposed method fused multiple biosignals at the input level to fully account for their interactions during model training (Pawłowski et al., 2023). In order to consider individuals’ physiological differences, we built a personalized model with optimized parameters and hyperparameters for each nurse. We utilized SSL to learn baseline representations for each subject from their large pool of unlabeled data (Liu et al., 2023). Our methodology was evaluated on a dataset that consists of the biosignal readings of nurses during the early stages of the COVID-19 pandemic.

## 2. Related Work

### 2.1 Time Series Prediction with 1D CNNs

Much research has been conducted on predicting individuals’ emotional and stress states based on physiological data using ML and deep learning methodologies (Gedam & Paul, 2021). Among ML models for processing time series signals, the 1D CNN is proven to be high-performing due to its ability to learn local features (Acharya et al., 2018). A concrete demonstration of the efficacy and high performance of the 1D CNN is demonstrated in the work of Santamaria-Granados et al. (2019), where a 1D deep CNN is applied to the AMIGOS dataset, which examines neurophysiological signals of individuals while they view emotional videos in different social settings (Miranda-Correa et al., 2021).

### 2.2 Multimodal Fusion and Machine Learning

Recent work in the field has highlighted the importance and potential of integrating multiple modalities within a single model (Jabeen et al., 2023), especially in the biomedical domain (Stahlschmidt et al., 2022). A number of physiological datasets incorporate diverse features such as the electrocardiogram (ECG), electrodermal activity (EDA), respiratory activity (RSP), and electroencephalogram (EEG) (Rim et al., 2020). The three primary multimodal data fusion techniques are input-level, feature-level, and decision-level fusion.

#### 2.2.1 Early Fusion

The early or input-level fusion method extracts and combines data before learning higher-level features deeper in the network to take into account the interaction between modalities throughout the network (Pawłowski et al., 2023). Despite its benefits, the early fusion process can be very intricate as raw modalities are likely to display heterogeneous characteristics (Liang et al., 2023).

#### 2.2.2 Intermediate Fusion

The intermediate fusion method, also known as feature-level fusion, provides flexibility in merging representations either at a single layer or through multiple layers during various stages of model building. Chen et al. (2022) generated a new dataset (n=52, features=ECG, EDA, RSP) and built end-to-end multimodal deep CNN structures that enable feature-level fusion. Fouladgar et al. (2022) developed CN-waterfall, a deep CNN for multimodal physiological effect detection that comprises base and general modules. In the base module, features were extracted separately, and in the general module, modalities were fused, considering uncorrelated and correlated modalities. Ross et al. (2019) applied intermediate fusion to EDA and ECG data for multimodal expertise classification in adaptive simulation using various ML algorithms.

#### 2.2.3 Late Fusion

The late fusion method, which fuses data after training individual models for each modality, is usually the simplest technique. According to Roheda et al. (2018), the Bayesian rule and Dempster-Shafer fusion are among the most commonly used decision-level fusion techniques. The researchers created a new decision-level fusion model for sensors that considers the task as a combination of events and takes into account the degree of correlation among features.

### 2.3 Personalized Patient-Specific Methods

Precision health formulates tailored healthcare solutions that align with individuals’ distinct characteristics, such as genetic makeup and environmental factors (Johnson et al., 2021). Following the shift in trend to precision health, machine learning research endeavors, particularly those involving biosignal data, have started to implement individualized learning approaches. Kiranyaz et al. (2016) built a patient-specific 1D deep CNN with ECG data from the MIT-BIH arrhythmia database, achieving high accuracy and computational efficiency. In a similar vein, Li et al. (2018) utilized ECG data to first develop a generic CNN (GCNN) by aggregating data from all patients. Then, they applied fine-tuning techniques to develop patient-specific Tuned Dedicated CNN and confirmed that TDCNN achieves higher accuracy than the baseline GCNN.

### 2.4 Self-Supervised Learning

Self-supervised learning (SSL) is a broad ML paradigm of learning meaningful features from unlabeled data, and the optimal SSL method depends on the specific data type and research objective. SSL has emerged as a promising approach in various domains, such as computer vision (Wang et al., 2017) and natural language processing (Weld et al., 2009). Its utilization in neural network pre-training is gaining popularity as it increases model performance using previously ignored unlabeled data. The framework has been actively applied to biosignal data, especially on mental state classifications (Bhatti et al., 2021; Sarkar & Etemad, 2022). Montero Quispe et al. (2022) reviewed the application of SSL in emotion recognition models and developed both self-supervised and regular supervised CNN models for the task. In broad terms, SSL can be categorized into contrastive, generative, and adversarial learning, among other methods (Liu et al., 2023).

#### 2.4.1 Contrastive Learning

Contrastive learning is an SSL method with an encoder trained to learn by contrasting similar and dissimilar data. Deldari et al. (2022) introduced COCOA (Cross mOdality COntrastive leArning), which learns by reducing the similarity among unrelated features. They tested COCOA on five datasets (UCIHAR, SLEEPEDF, Opportunity, PAMAP2, WESAD) and demonstrated its superiority over baseline fine-tuned encoders. Kiyasseh et al. (2021) developed CLOCS (Contrastive Learning of Cardiac Signals), a self-supervised pre-training technique that encourages the similarity of representations across space, time, and patients and produces individualized representations.

#### 2.4.2 Generative Learning

Generative models reconstruct the input received in an encoder using a decoder, and they include auto-regressive models, auto-encoding models, and hybrid generative models. Chen et al. (2022) built MAEEG (Masked Auto-encoder for EEG Representation Learning), a specific generative model trained to reconstruct the hidden EEG attributes. The researchers used MAEEG to pre-train the sleep EEG dataset, increasing the accuracy of the sleep stage classification model by 5%.

#### 2.4.3 Adversarial Learning

Adversarial or generative-contrastive learning models are trained using an approach where the encoder and decoder collaborate to create adversarial data, while a discriminator distinguishes between adversarial samples and real data. Adversarial learning has found significant applications in biosignal data augmentation. For instance, Haradal et al. (2018) proposed a generative adversarial network with each neural network performing similarly to recurrent neural networks (RNN) and long short-term memory (LSTM). They employed their proposed method to augment ECG and EEG data, improving biosignal classification performance.

## 3. Methods

### 3.1 Dataset Description

We trained SIM-CNN using the Nature Scientific Data dataset release entitled “A multimodal sensor dataset for continuous stress detection of nurses in a hospital” (Hosseini et al., 2021). The data collection occurred in three phases in 2020 during the beginning of the COVID-19 pandemic. Nurses in their regular shifts were each given an Empatica E4 wristband, a wearable device that monitors six biometric signals, namely electrodermal activity, heart rate, skin temperature, interbeat interval, blood volume pulse, and accelerometer data. The E4 wristband transmitted the collected data to the nurse’s smartphone, and the smartphone forwarded it to the analytics server. Stress detection was performed in near real-time to identify potential intervals of stress through a Random Forest (RF) model that was trained, tested, and validated on the AffectiveRoad dataset (Haouij et al., 2018).

Surveys were administered for each nurse to label no-stress, medium-stress, or high-stress for each interval where the RF model detected stress. Even though the system was able to detect stress in real-time, the app collected survey responses at the end of each shift for every nurse to minimize interference. The nurses were requested to report any undetected stress intervals via their wristbands or survey responses, but none used this option. Therefore, the supposedly no-stress intervals could not be validated, leading to a substantial amount of unlabeled data. 15 nurses completed the study, and around 1,250 hours of data was collected, with 171 hours identified as stressful by the RF model and 83 of those hours labeled with stress levels by the nurses. To ensure anonymity and maintain the separation of individual nurse data and survey responses, a distinct identifier was assigned to each nurse.

### 3.2 Data Preprocessing

Figure 1 depicts the data preprocessing pipeline implemented to create the labeled dataset for each nurse. Initially, the biometric signal data of each modality for every nurse was extracted. This data was filtered to retain only the parts with matching non-null survey results, i.e., the parts that the RF model predicted as stressful and the nurses labeled with stress levels. The corresponding stress labels were extracted from the survey results and were saved along with the filtered data. We disregarded the survey results that lacked the corresponding raw data, which likely arose due to the flaws in the dataset.

**Figure 1.**
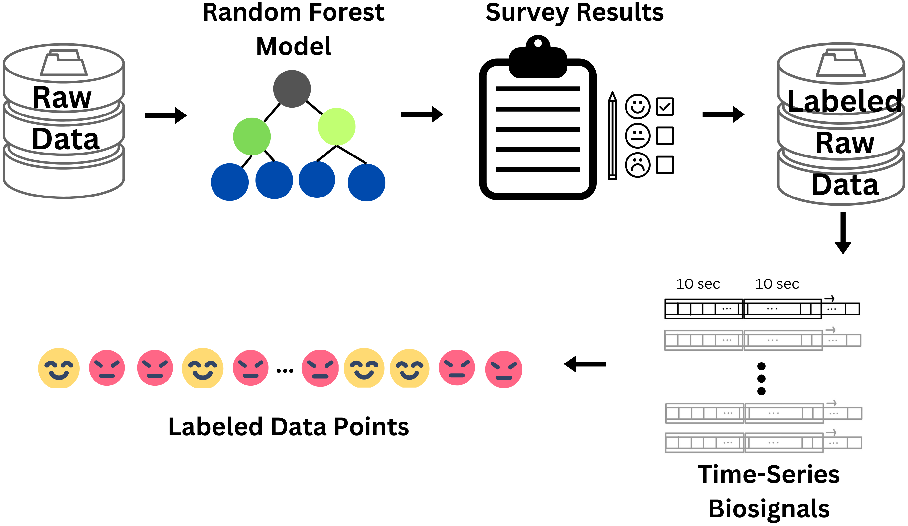
Labeled dataset creation and data preprocessing steps performed for each nurse. The unlabeled dataset follows a similar pipeline but excludes survey results.

There was a discrepancy between the epoch timestamps of the dataset and the start and end times of the stress-detected intervals that were in human-readable time rounded to minutes. The less-than-60-second difference caused by rounding resulted in some of the valid stress-detected intervals not being fully contained in the raw data. To eliminate this error, a minute was added to the start time and subtracted from the end time of each interval.

A 10-second window was constructed such that each sample would contain biosignal data recorded for 10 seconds. This data point extraction procedure was applied to both the filtered and unfiltered signals for each nurse to generate the labeled and unlabeled samples, respectively. The window size was chosen so that there was a sufficient number of data points while each data point was not excessively small. The interbeat interval was excluded from the analysis due to missing data, and the x, y, and z axes of the accelerometer were treated as three distinct modalities, resulting in seven modalities in total.

Each modality was recorded at varying frequencies, ranging from 1 Hz to 64 Hz. To ensure temporal alignment, the time series data for all modalities was interpolated or downsampled to a consistent frequency of 8 Hz. Graphs of both the original data and downsampled data were plotted to verify that no significant information was lost during this process. The downsampled data for each modality was then concatenated along the feature dimension to generate each nurse’s multimodal dataset.

The labeled train, validation, and test sets were created for each nurse so that their class distributions were identical to that of the entire data of the nurse. Shuffling was not performed, as it may have resulted in data leakage due to the nature of time series data. The task originally involved three classes: no-stress, medium-stress, and high-stress. However, due to the lack of medium-stress data and the potential bias associated with differentiating between medium-stress and high-stress, medium-stress data was completely removed. The task was therefore treated as the binary classification between stress and no-stress.

To address the class imbalance, the data points in the minority class were upsampled to match the number of data points in the majority class. The minority class was the nostress class for most nurses, as the labeled data consisted of intervals where the original RF model detected stress. Two nurses with total absence of no-stress data were eliminated.

### 3.3 Model Architecture

For each nurse, a baseline multimodal CNN model was built with the architecture presented in Figure 2 to predict their stress level. The 1D CNN architecture was selected due to its unidirectional kernels that generally lead to performant time series models. Early fusion was implemented to concatenate all modalities over the feature dimension at the input level so the model could capture the interactions between modalities while training.

**Figure 2.**
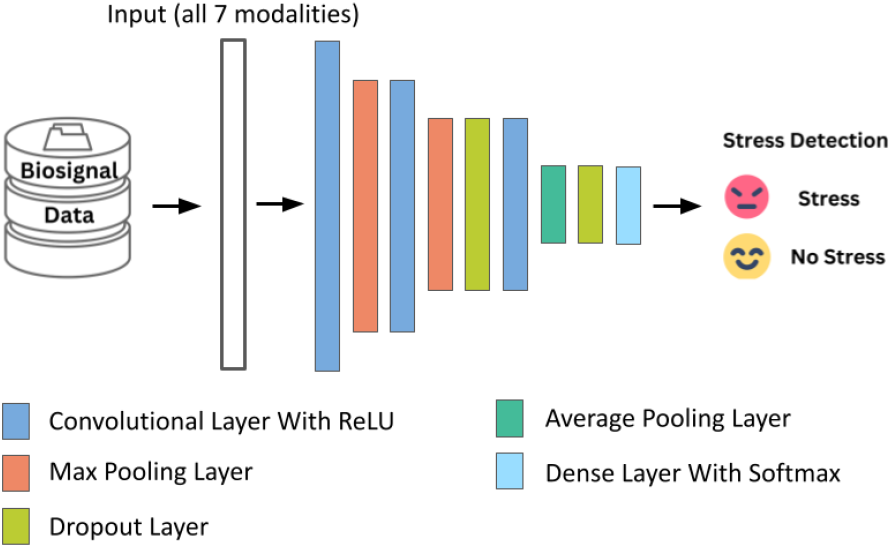
Illustration of the baseline individualized multimodal CNN model architecture that predicts stress using biosignal data.

As depicted in Figure 2, the proposed model comprises convolutional layers, max pooling layers, and dropout layers, with an average pooling layer and a dense layer added at the end for prediction. Each convolutional layer has the ReLU activation function that addresses the need for non-linearity without causing issues with gradients and backpropagation. Max pooling and average pooling layers help reduce overfitting while downsampling the spatial dimension. Dropout layers also prevent overfitting by randomly ignoring some of the outputs of the previous layers. The final dense layer employs the softmax activation function to produce a probability distribution over the two classes: stress and no-stress.

#### 3.3.1 Hyperparameter Search and Threshold Optimization

The initial hyperparameter search was conducted on Nurse E4, exploring variations in feature mappings (ranging from 16 to 128), kernel size (5 or 10), batch size (16, 32, or 64), learning rate (1e-4 or 1e-5), and training epochs (25, 50, or 75). The max pooling layers contained a pool size of 2, and the dropout layers had a rate of 0.3. The Adam optimizer was used to train the models. The search concluded that the optimal model architecture has a kernel size of 5 and 16 feature mappings for the first two convolutional layers and 32 feature mappings for the last convolution layer. Building upon this baseline architecture, we optimized the training hyperparameters for each nurse – namely the batch size, learning rate, and the number of training epochs – by selecting the model with the highest validation set average AUC (area under the receiver operating characteristic curve) for the last five epochs. A probability threshold is used in a binary classification model to determine the predicted class. If the probability of the stress class produced by the softmax activation function is above the threshold, our model predicts stress; otherwise, the model predicts no-stress. For each personalized model, the probability threshold that yielded the highest validation set F1 score was selected.

### 3.4 SIM-CNN

#### 3.4.1 Self-Supervised Pre-Training

To harness the vast amount of unlabeled data, a self-supervised pre-training task was employed to learn the various patterns in the biosignals and the interplay between different modalities. In particular, as illustrated in Figure 3, the model was pre-trained on the unlabeled data to predict the values of the seven features for the next time step given the data for the preceding 10-second window. This pretraining task, called next-step time series forecasting, lays the foundation that would be valuable during the subsequent fine-tuning of the model for stress recognition.

**Figure 3.**
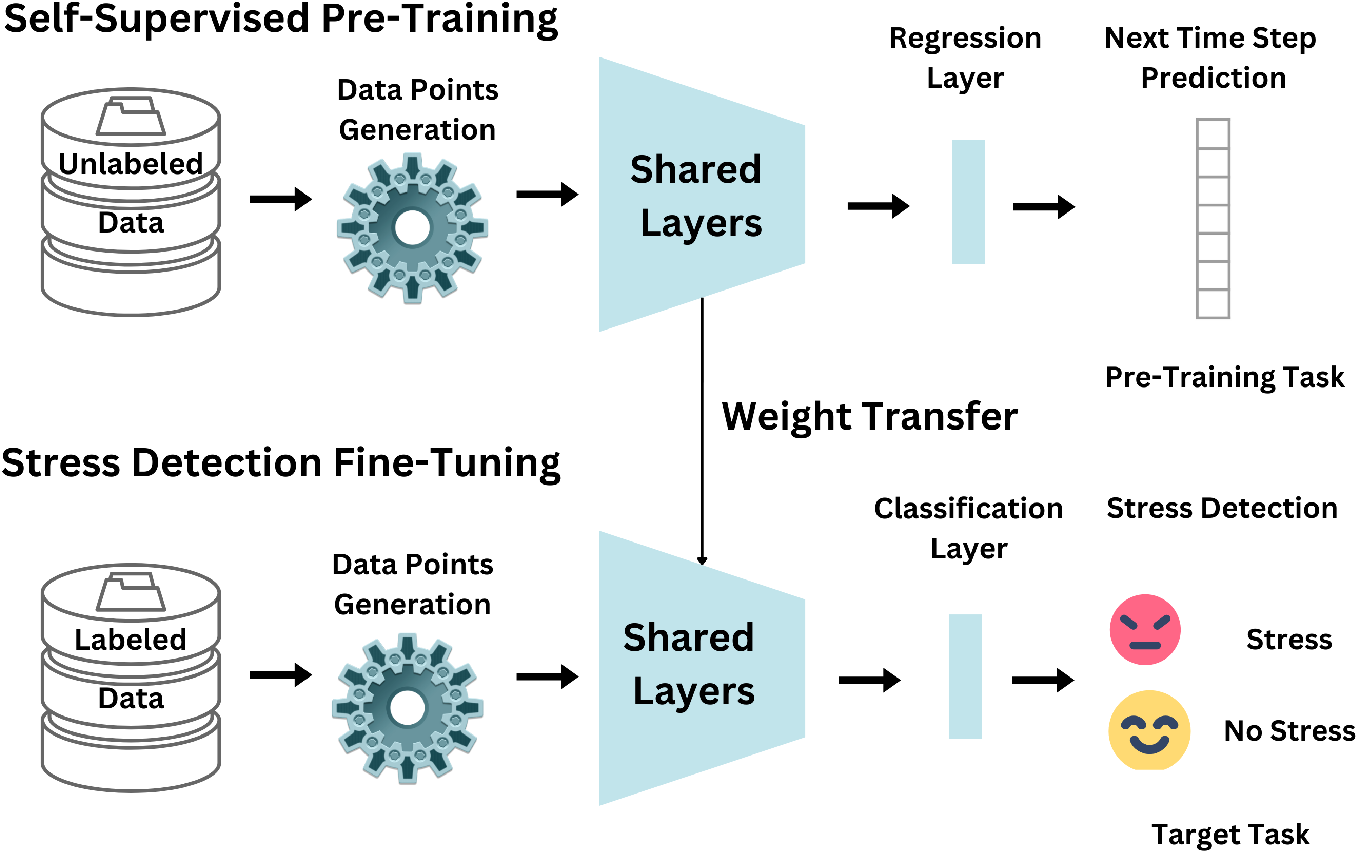
Overview of the architecture of self-supervised individualized multimodal convolutional neural networks (SIM-CNN) pre-trained on the unlabeled data for next-step time series forecasting and fine-tuned on the labeled data for stress prediction.

#### 3.4.2 Fine-Tuning for Stress Recognition

The model was then fine-tuned on the labeled data for the supervised stress-prediction task. A new dense layer that generates a probability distribution for classification replaced the regression layer. Compared to the baseline model, where the weights were initialized randomly, the fine-tuned model’s initial weights were obtained from the self-supervised pre-training. This approach equips the model with prior knowledge about the data before fine-tuning on the labeled data, which results in improved performance over the baseline model.

#### 3.4.3 Hyperparameter Search and Threshold Optimization

The identical architecture as the baseline CNN shown in Figure 2 was used in SIM-CNN for each nurse to properly assess the effect of self-supervised pre-training. We searched over the pre-training hyperparameters for each individual, selecting the best-performing model and optimizing the threshold with the same methods as the baseline CNN.

## 4. Results

### 4.1 Evaluation Metrics

The metrics used to evaluate our model performance are AUC, F1 score, and accuracy. In this context, “positive” refers to the prediction of stress, and “negative” signifies otherwise. F1 score is the harmonic mean of positive predictive value (PPV) and true positive rate (TPR). PPV, or precision, represents the proportion of correctly-predicted positives among true positives and false positives. TPR, or recall, quantifies the proportion of the correctly-predicted positives among true positives and false negatives. AUC calculates the area under the TPR and false positive rate (FPR) at various thresholds, quantifying the model’s ability to distinguish between the two classes at each threshold. FPR quantifies the proportion of the falsely-predicted positives among true negatives and false positives.

### 4.2 Model Performance

In order to establish performance baselines for our models, we trained Support Vector Machines (SVMs). According to the metrics presented in Table 1, there is an overall improvement in the average AUC and accuracy from SVM to the baseline CNN, as well as from the baseline CNN to SIM-CNN. The average F1 score remains consistent across the different models.

**Table 1.** Average of threshold and metrics on the test set across all 13 nurses.

| MODEL | AUC | F1 SCORE | ACCURACY |
| --- | --- | --- | --- |
| SVM | 55.40 | 76.99 | 71.70 |
| CNN | 58.47 | 75.11 | 75.72 |
| SIM-CNN | 60.96 | 75.22 | 79.65 |

SIM-CNN overall outperforms the baseline CNN, with the average AUC improving from 58.47% to 60.96% and accuracy from 75.72% to 79.65%. Compared to SVM, SIM-CNN shows a 5.56% increase in the average AUC and a 7.95% increase in the average accuracy; the baseline CNN shows a 3.07% increase in the average AUC and 4.02% increase in the average accuracy.

The specific metrics and threshold for each nurse can be observed in Table 2, which provides a detailed view of SIM-CNN’s performance. When observed on an individual basis, each SIM-CNN model demonstrates significant improvements from the baseline CNN in AUC, F1 score, and accuracy, improving up to 24.74%, 11.71%, and 15.8%, respectively. Notably, F5’s SIM-CNN achieves 100% for the AUC and over 99% for the F1 score and accuracy.

**Table 2.** Threshold and performance metrics on the test set for each nurse.

| NURSE | MODEL | THRESHOLD | AUC | F1 SCORE | ACCURACY |
| --- | --- | --- | --- | --- | --- |
| E4 | SVM | 0.05 | 25.03 | 89.45 | 80.95 |
|  | CNN | 0.25 | 37.18 | 70.41 | 54.33 |
|  | SIM-CNN | 0.42 | 61.92 | 82.12 | 70.13 |
| 83 | SVM | 0.01 | 53.91 | 90.47 | 82.59 |
|  | CNN | 0.01 | 52.10 | 91.48 | 84.62 |
|  | SIM-CNN | 0.01 | 45.98 | 90.47 | 82.59 |
| 94 | SVM | 0.07 | 36.82 | 38.30 | 37.86 |
|  | CNN | 0.16 | 36.37 | 33.08 | 36.43 |
|  | SIM-CNN | 0.55 | 50.36 | 12.82 | 51.43 |
| BG | SVM | 0.01 | 65.05 | 98.66 | 97.35 |
|  | CNN | 0.01 | 67.39 | 98.39 | 96.83 |
|  | SIM-CNN | 0.01 | 50.54 | 98.66 | 97.35 |
| DF | SVM | 0.01 | 67.33 | 95.80 | 91.93 |
|  | CNN | 0.01 | 67.28 | 92.57 | 86.32 |
|  | SIM-CNN | 0.01 | 71.13 | 95.97 | 92.28 |
| F5 | SVM | 0.08 | 98.10 | 96.91 | 94.05 |
|  | CNN | 0.05 | 100.00 | 99.06 | 98.21 |
|  | SIM-CNN | 0.08 | 100.00 | 99.68 | 99.40 |
| 6B | SVM | 0.88 | 64.27 | 63.16 | 51.23 |
|  | CNN | 0.68 | 66.30 | 75.07 | 63.52 |
|  | SIM-CNN | 0.57 | 71.22 | 70.06 | 59.02 |
| 6D | SVM | 0.01 | 32.21 | 24.34 | 23.12 |
|  | CNN | 0.71 | 56.83 | 11.76 | 83.87 |
|  | SIM-CNN | 0.94 | 26.97 | 6.25 | 83.87 |
| 7A | SVM | 0.01 | 44.20 | 97.52 | 95.16 |
|  | CNN | 0.01 | 49.46 | 95.80 | 91.94 |
|  | SIM-CNN | 0.01 | 59.97 | 97.00 | 94.19 |
| 7E | SVM | 0.05 | 83.53 | 69.92 | 53.75 |
|  | CNN | 0.39 | 69.26 | 74.75 | 68.75 |
|  | SIM-CNN | 0.27 | 82.28 | 80.00 | 75.00 |
| 8B | SVM | 0.01 | 24.05 | 80.36 | 67.16 |
|  | CNN | 0.01 | 11.72 | 80.36 | 67.16 |
|  | SIM-CNN | 0.01 | 20.52 | 80.36 | 67.16 |
| 15 | SVM | 0.01 | 58.92 | 97.38 | 94.90 |
|  | CNN | 0.01 | 76.67 | 97.38 | 94.90 |
|  | SIM-CNN | 0.01 | 78.71 | 97.38 | 94.90 |
| 5C | SVM | 0.56 | 66.80 | 58.54 | 62.01 |
|  | CNN | 0.71 | 69.53 | 56.32 | 57.54 |
|  | SIM-CNN | 0.69 | 72.83 | 67.05 | 68.16 |

SIM-CNN employs a novel method that integrates multimodality, self-supervised learning, and personalization in stress prediction on a new dataset that has not been previously used. Consequently, direct comparisons between SIM-CNN and prior research are not possible.

The average metrics across the nurses demonstrate SIM-CNN’s superiority in stress prediction and the improvement of the baseline CNN from the SVM. However, the metrics for some nurses, namely 83, BG, 6D, 7E, and 8B, deviate from this trend. Specifically, 83, 6D, and 8B exhibit unusually low AUCs that do not improve even with the implementation of SIM-CNN. The thresholds, combined with each nurse’s dataset size and class distribution shown in Table 3, help explain these results. We emphasize that all three models for 83, BG, and 8B have a threshold of 0.01, meaning that they are not successful in learning to predict the no-stress class and mostly or exclusively predict the stress class. This suggests the presence of dataset-related issues specific to these nurses.

**Table 3.** Summary statistics of the dataset size and class distribution of each nurse.

| NURSE | STRESS PERCENT | NO-STRESS TRAIN | NO-STRESS VAL | LABELED | UNLABELED |
| --- | --- | --- | --- | --- | --- |
| E4 | 80.88 | 703 | 89 | 4607 | 41805 |
| 83 | 82.68 | 340 | 43 | 2460 | 53897 |
| 94 | 29.74 | 782 | 98 | 1392 | 28943 |
| BG | 97.45 | 38 | 5 | 1884 | 27325 |
| DF | 92.81 | 162 | 21 | 2838 | 45956 |
| F5 | 94.27 | 76 | 10 | 1674 | 25032 |
| 6B | 86.17 | 268 | 34 | 2430 | 20903 |
| 6D | 14.94 | 1256 | 158 | 1848 | 8619 |
| 7A | 95.34 | 114 | 15 | 3090 | 28529 |
| 7E | 53.79 | 292 | 37 | 792 | 18236 |
| 8B | 67.35 | 344 | 44 | 1323 | 9788 |
| 15 | 95.65 | 32 | 5 | 966 | 26197 |
| 5C | 45.79 | 772 | 97 | 1782 | 35743 |

Delving deeper into the individual datasets of the nurses whose models are underperformant, we consistently observe extreme class imbalance and limited dataset size. BG has the most imbalanced dataset, with 97.45% of the labeled data being from the stress class and containing only five samples of the no-stress class in its validation set. 8B has very small unlabeled and labeled datasets, preventing the models from learning to perform the complex stress-prediction task. Similarly, 7E’s labeled dataset has the smallest size. Furthermore, 6D not only has the smallest unlabeled dataset but also has the lowest stress percent of 14.94%. This means that the RF model had a precision of 14.94% for this nurse, suggesting a significant amount of noise in the dataset. Although the specific cause of 83’s low performance is not evident from Table 3, the threshold of 0.01 indicates issues within the dataset.

## 5. Discussion

We aimed to build high-performing stress-prediction models and to determine the effect of SSL on model personalization. To achieve this, we constructed a baseline individualized multimodal CNN and an identical model trained with SSL (SIM-CNN). By comparing the personalized models with each other and with the baseline SVMs, we showed that, on average, SIM-CNN outperforms the baseline CNN, and the baseline CNN outperforms SVM. For the few nurses whose metrics do not follow this pattern, we revealed that imbalanced datasets and the lack of data create a challenge for developing accurate, personalized models for these nurses.

### 5.1 Limitations and Future Work

Although SIM-CNN improves the average AUC and accuracy, further investigation of the data from certain nurses whose model metrics differ from the expected pattern reveals the intrinsic limitations of precision health using in-the-wild biosignal data. Due to its inherent subjectivity and complexity, stress prediction solely based on biosignals is a difficult task for computational approaches. The real-world dataset that we used unavoidably contains biases and class imbalance, as all labeled data was collected from the time intervals that a classifier originally predicted as stressful. In addition to sampling bias, the nurses might have been biased when self-labeling the intervals since no external verification was conducted. Moreover, the nurses had to label the entire stress-detected interval with one stress level, even though it often lasted over 30 minutes and sometimes over 2 hours, resulting in a lot of noise in the labeled data.

Combined with the individualized aspect of each model, which entails relatively smaller datasets that are more susceptible to noise, bias, and class imbalance, these complications explain why some nurses’ metrics are low or distinct from the overall pattern of improvement across the three models. These issues can be mostly addressed by using larger labeled and unlabeled datasets. Collecting unlabeled biosignal data is feasible, especially in medical settings where subjects can wear devices that continuously monitor their biosignals. However, generating a substantial amount of manually-labeled data is time-consuming and costly, particularly within the healthcare domain.

To evaluate SIM-CNN’s performance in the absence of these complications, it would be beneficial to train the model on a cleaner dataset with more labeled data. Such datasets can be obtained by alleviating the reliance on manual annotation, as it would reduce bias and result in more labeled data. This could also involve using data collected from subjects in professions outside of the healthcare domain since healthcare workers’ data carries unique biases and noise due to irregular work shifts, the need to minimize interruptions during work hours, and other environmental factors.

There are two potential avenues to acquire such datasets. The first approach entails the presence of on-site observers who can identify stress intervals. Although this component was part of the initial design of the study that generated our dataset, it could not be realized due to the social distancing requirements imposed by the pandemic. The second approach is to gather data from individuals in professions where stress intervals can be objectively identified through external factors. For instance, one method involves the collection of pilots’ biosignals in conjunction with data on flight start and end times, as well as timestamps corresponding to any warnings or unforeseen issues encountered during flights. Another method would be to collect biosignal data from service workers while concurrently recording the number of clients they attended to within each time interval. Furthermore, occupations characterized by consistent daily deadlines, such as employees involved in daily news production, present intriguing possibilities.

SIM-CNN predicts stress at a performance surpassing that of the baseline CNNs and SVMs, demonstrating its potential for effective utilization across various domains. Notably, SIM-CNN holds promise for tasks in the field of affective computing that share commonalities with stress prediction, such as emotion classification. There is also potential for transfer learning of SIM-CNN weights for stress recognition for these related tasks. Furthermore, SIM-CNN extends its application beyond mental states to the diagnosis of physical health disorders, such as epilepsy, which exhibits a strong association with physiological signals. SIM-CNN is capable of adapting the number and types of signals based on the specific task requirements, enhancing its versatility and applicability.

SIM-CNN, which implements the next-step time series forecasting, does not appear to achieve high performance without sufficient labeled data. A potential future study could involve investigating other types of SSL that could enhance model performance with limited labeled data. We could also pre-train the models using data from all subjects or explore other ways to effectively combine and utilize other subjects’ data before personalizing the models. Furthermore, an updated version of SIM-CNN could be deployed into an end-to-end system designed for real-time stress recognition using a wearable device and an accompanying application.

The model personalization approach we introduce can be applied to a variety of precision health tasks beyond stress prediction. There are myriad applications in digital health where vast streams of unlabeled data are recorded with a few corresponding user-provided labels, such as continuous blood glucose monitoring systems for diabetes management (Makroum et al., 2022; Plis et al., 2014; Kavakiotis et al., 2017), digital autism therapeutics (Daniels et al., 2018; Voss et al., 2019; Washington et al., 2020; 2017; Kalantarian et al., 2019), and audio-based continuous monitoring of lung health (Xu et al., 2021; Vatanparvar et al., 2020). We encourage the precision health research community to explore the application of the individualized learning framework described here to these other healthcare domains.

## 6. Conclusion

SIM-CNN is a novel personalized learning framework that integrates multiple physiological modalities while leveraging unlabeled data through self-supervised learning. This innovative approach entails constructing models with individualized parameters and hyperparameters for precision health. Such personalization of models is necessary to handle the inherent subjectivity and heterogeneity of stress data and of complex social human data more broadly (Washington et al., 2020; 2022; Washington & Wall, 2023). SIM-CNN’s performance increase in individual and average metrics underscores its potential applications in mitigating various medical complications. This performance is particularly noteworthy considering that stress encompasses multidimensional aspects beyond those depicted in biosignals, coupled with the fact that the real-world dataset that we used to evaluate SIM-CNN contains high variability. The robust performance of SIM-CNN affirms the effectiveness of using SSL to enhance stress-recognition models, thereby offering a potential solution to the intrinsic complexity of the underlying prediction task.

## Data Availability

We used a publicly available dataset published in Nature Scientific Data.

